# The PROVIDE Study: Primary care assessment of the ROche IPDM tools for Validation and Implementation in Diabetes management and Evaluation

**DOI:** 10.1101/2023.12.23.23300283

**Authors:** Simran Roshan, Elena Hunter, Claire Marriott, Ross Carruthers, Barnaby Poulton, Linda Gibson, Jeremy Cummin, Fahad Rizvi, Rusi Jaspal, Philippe B. Wilson

## Abstract

**Introduction:** The adoption of digital tools in diabetes care has transformed the way people with diabetes (PwD) and healthcare professionals (HCPs) manage the condition. Digital tools allow for remote management, improved accuracy in blood glucose readings, and enhanced education for patients. This paper presents the results of a 12-month service evaluation of the Roche Diabetes Care Platform, a diabetes data management platform that was implemented into a primary care network (PCN). The aim of the implementation of the RocheDiabetes Care Platform was to address the current challenges faced by clinicians and PwD in diabetes care, such as reducing complications, improving diabetes education, and the delivery of health care to PwD. Semi-structured interviews were conducted virtually to capture the views of HCPs on their experiences with these digital tools.

**Methodology:** Interviews were conducted with individuals in a range of roles, intrinsic to the diabetes service management in the Leicester PCN, at each stage of implementation. This included pre-implementation, implementation, and post-implementation phases. The data was analysed using thematic analysis.

**Results:** The use of the RocheDiabetes Care Platform in the Leicester PCN has advanced HCPs’ management of PwD, demonstrably improving patient outcomes and enhancing interactions. There were two themes identified in the pre-implementation stage, including pre-appointment activities and challenges, and appointment activities and challenges. A further six themes were identified in the post-implementation results, including the optimisation of appointments, time and cost savings, enhanced engagement with PwD, the centralisation of resources, challenges in the PCN’s pathway of care that the RocheDiabetes Care Platform addressed and the future vision of the PCN with the RocheDiabetes Care Platform. Overall, available and accessible data helped to reduce the number of appointments efficiency. Additionally, where PwD were onboarded onto the RocheDiabetes Care Platform, and there was no specific need for the PwD to attend the practice, the number of face-to-face appointments was reduced, improving overall clinic efficiency. Over the period of the evaluation there were no (0) Did Not Attend (DNAs).

## 1. Introduction

Currently, 4.3 million people are living with diabetes in the UK. Diabetes UK estimates that there could be more than 2.4 million people at a high risk of developing type 2 diabetes.^1^ Diabetes is a chronic and increasingly prevalent condition that requires the continual management of blood sugar levels, diet, and exercise. Self-management plays a large role in improving glycemic control and reducing the risk of complications associated with the condition.^2^.

Integrated Personalised Diabetes Management (iPDM) is an approach that leverages connected digital tools to bring insights to people living with diabetes and their HCPs. This includes digital software for diabetes data management for HCPs and connected blood glucose meters for the self-monitoring of blood glucose (SMBG), for people who live with diabetes.

The recent move towards digitisation in diabetes care has accelerated the potential of this personalised care; using digital tools to capture diabetes data and translate it into meaningful insights improving the timeliness of therapy interventions, quality of life and outcomes for people with diabetes.^3^

Using this approach can increase patient and HCP collaboration in consultations.^4^ This new era of diabetes care aims to improve both patient quality of life and healthcare professional efficiency. Consequently, there have been calls for PCNs to adopt digital interventions into their pathway of care, to better support those living with diabetes and address the growing crisis.^5^

Furthermore, there is the potential for digital tools to address the challenges faced by HCPs and PwD in primary care. HCPs in primary care can face time constraints and limited resources when managing their PwD.^6^ Alongside this, the lasting effects of the COVID-19 pandemic remain and HCPs may be overworked in performing the backlog of routine diabetes care for PwD.^7^

The RocheDiabetes Care Platform is a diabetes management solution that organises personal diabetes data, intending to improve decision-making and delivery of care. The platform supports multiple devices, providing an interactive dashboard, status cards, and pattern detection, as well as graphs and reports, which helps to facilitate remote monitoring of PwD. The chosen PCN for the PROVIDE study embodies general practice sites across Leicester, incorporating 34,185 patients, 3,127 of whom have been diagnosed with type 2 diabetes. The chosen location for the service evaluation was Leicester, which had a substantial proportion of individuals with diabetes; 9.7% of Leicester’s population live with diabetes compared with the national average of 7.1 %8. There were 3 phases to this service evaluation: pre-implementation, implementation, and post-implementation. The perceptions of HCPs and the shift in their approach toward diabetes management were documented throughout this process. Interviews have been conducted across a range of diabetes clinical and support roles within the PCN.

To examine the effects of digital tools on diabetes management, a service evaluation was undertaken using the RocheDiabetes Care Platform and associated tools.

### Overview of the Roche Diabetes Care tools

As part of this service evaluation, HCPs were trained on the RocheDiabetes Care Platform’s functions. All patients were using the Accu-ChekⓇ Instant blood glucose meter, which sends blood glucose data directly to the mySugrⓇ app and connects to the RocheDiabetes Care Platform. Subsequently, PwD were onboarded onto the platform and taught to use the mySugr app. The descriptions of the platform and the app are provided below.

#### The RocheDiabetes Care Platform

The RocheDiabetes Care Platform aims to support HCPs with their population of PwD. The platform allows for the remote monitoring of diabetes data. The RocheDiabetes Care Platform aims to use graphs, charts, and statistics to present diabetes data in an easy to assess and standardised manner.

The RocheDiabetes Care Platform’s functions include an interactive dashboard, with information on blood glucose levels through the presentation of graphs and reports. These graphs display vital diabetes information regarding the time of day when blood glucose levels spike and fall. Status cards are all included in the platform and provide a summary of PwD’s glycaemic information. Moreover, the RocheDiabetes Care Platform detects patterns in patient data. The platform detects different patterns that can be set by the HCP on a patient level including hyperglycaemia, hypoglycaemia and treatment adherence. The digital tool can provide visual indicators for up to 20 different glucose patterns based on customisable therapy guidelines.^9^ (Figures 1 and 2).

**Figure 1.**
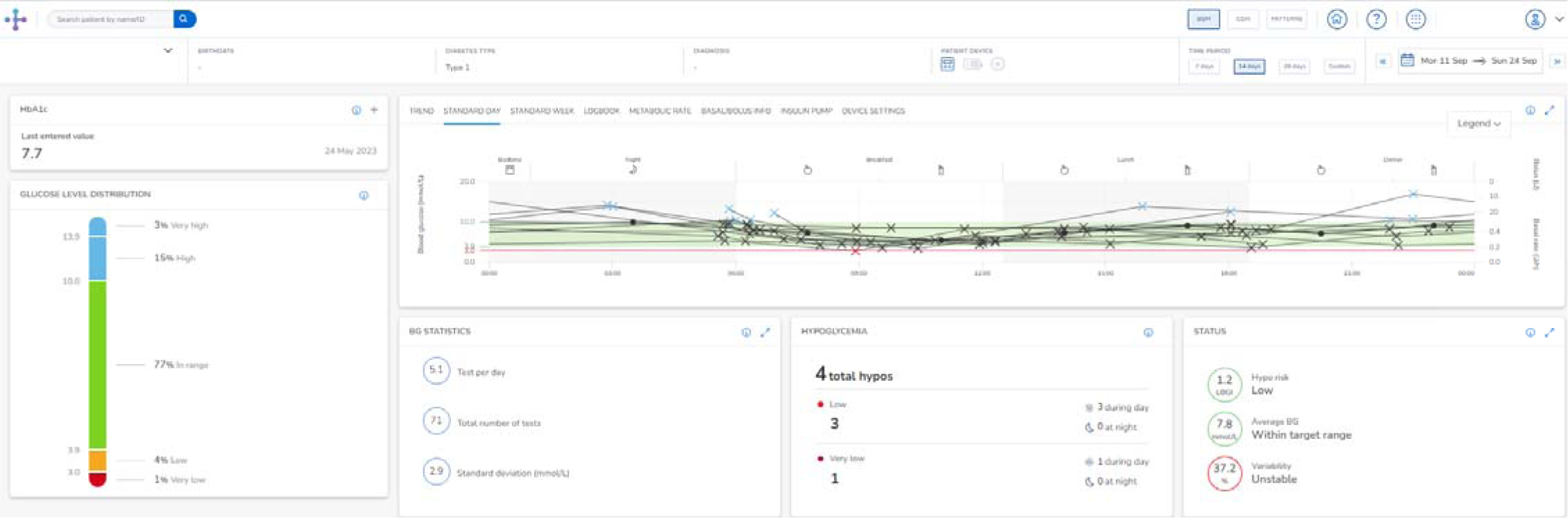
The RocheDiabetes Care Platform’s Interactive Dashboard, displaying patient status cards and pattern detection in patient blood glucose levels. The data presented within this figure represents a fictional patient created to display the functionality of the software.

**Figure 2.**
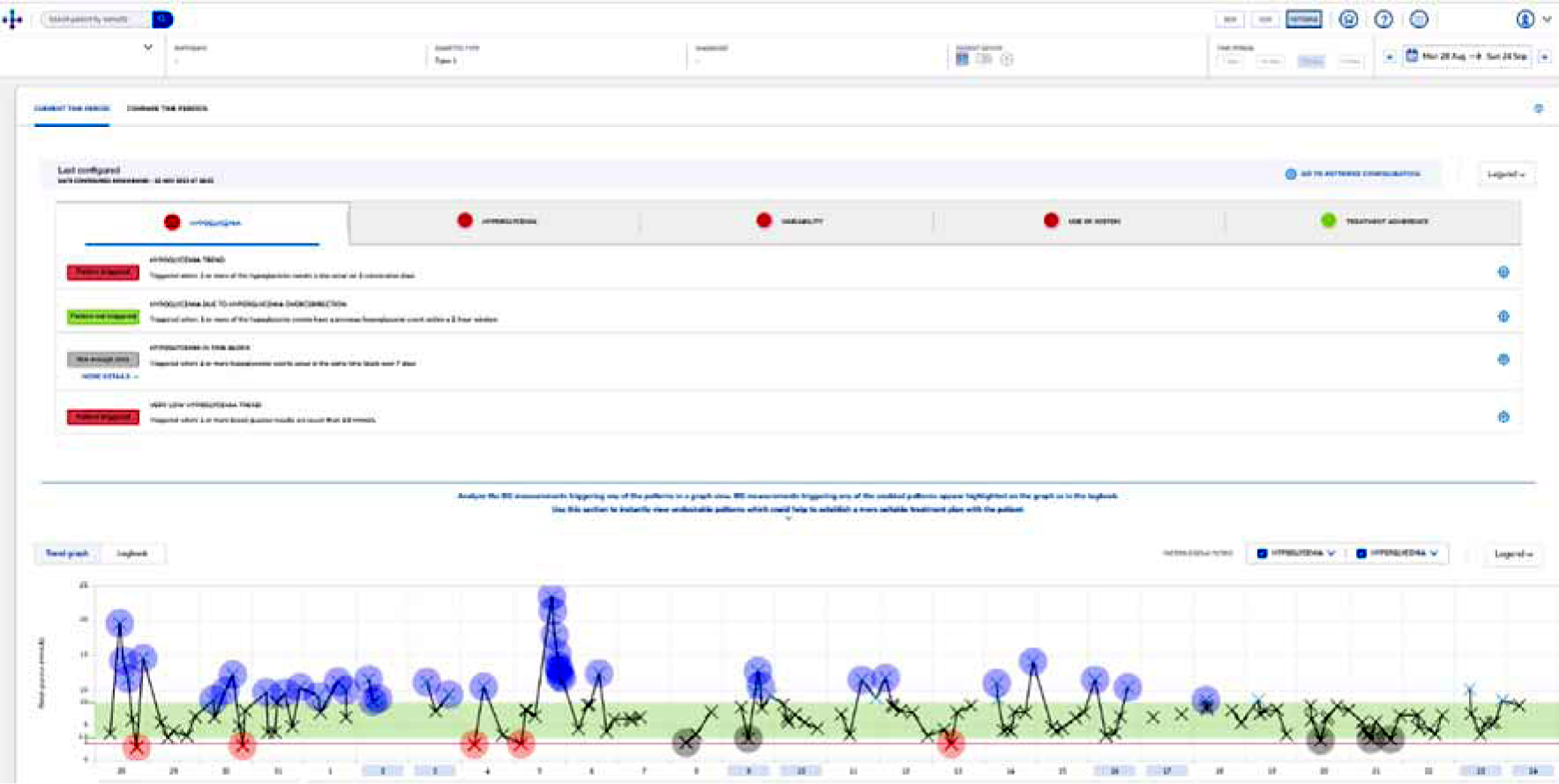
An example of the display of glucose readings, generated by the RocheDiabetes Care Platform.

The functions of the RocheDiabetes Care Platform aim to alleviate some of the challenges faced by HCPs in diabetes management, such as the lack of data provided by PwD for informed decision-making.^1^0

The RocheDiabetes Care Platform was the digital tool implemented for the use of HCPs within this service evaluation. The PwD did not have access to the RocheDiabetes Care Platform however the platform was displayed and discussed in onboarding appointments with HCPs in order to explain the relation between the RocheDiabetes Care Platform and the mySugr app. The PwD used the mySugr app to record and monitor their diabetes. The functions of this app are described below.

#### The mySugr app

The mySugr app is a tool for the use of PwD. The app stores diabetes data, including information from blood glucose meters, as well as manually entered data and provides PwD with an overview of their diabetes management (Figure 3). The connection between blood glucose meters and the mySugr app facilitates the automatic importing of blood sugar readings into the app, with the aim of decreasing the need to constantly upload data manually. Furthermore, PwD are able to track eating habits and fitness data using the app by connecting tools such as Google Fit or Apple Health (Figure 4). Therefore, PwD can share their data with HCPs through this integration of the mySugr app and the RocheDiabetes Care Platform.

**Figure 3.**
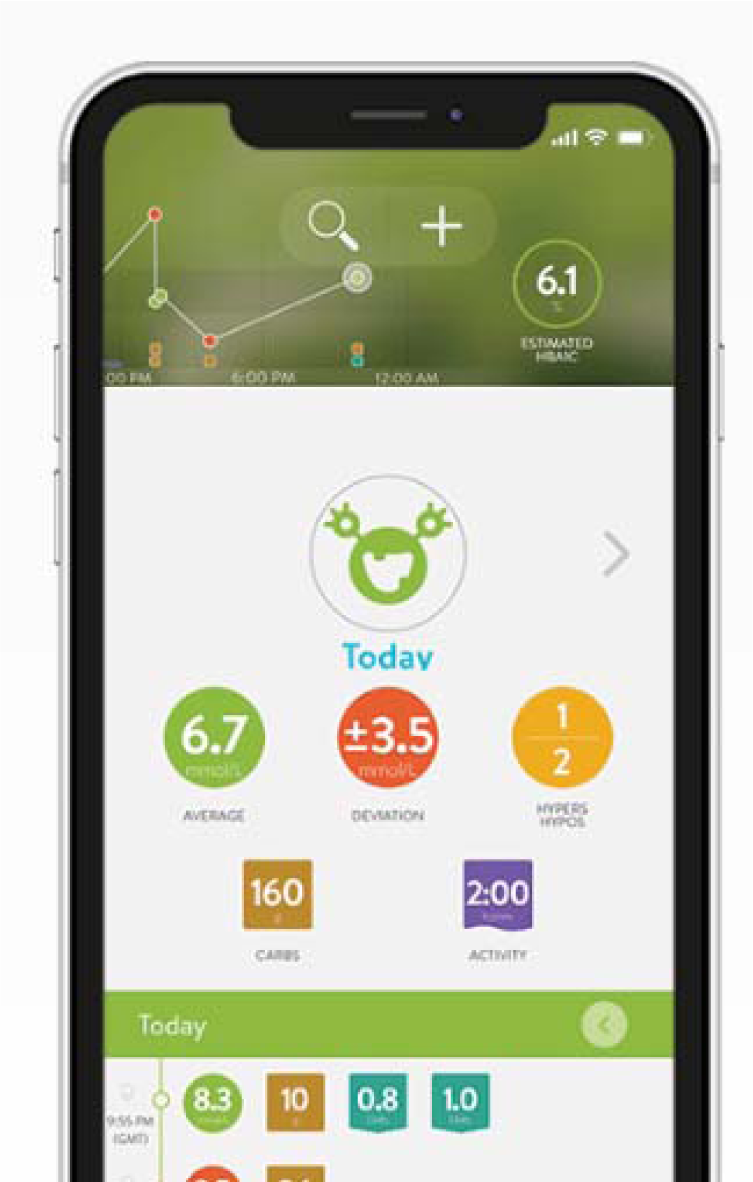
The mySugr app home screen for patients contains displays of average range, deviation and incidents of Hyper and Hypos.

**Figure 4.** The mySugr app logging page, where blood sugar readings, fitness, insulin and meals can be logged.

The RocheDiabetes Care Platform was implemented into the Leicester PCN. PwD in the PCN also downloaded the mySugr app onto their mobile phones. The combined use of digital tools contributed to the approach of iPDM.

The aim of this service evaluation was to capture the transformation in diabetes care pathways facilitated by the utilisation of the Roche Diabetes Care tools. Furthermore, this service evaluation examined the impact of iPDM within a PCN, using the RocheDiabetes Care Platform and the mySugr app.

## Methodology

To evaluate the digital platform, the RocheDiabetes Care Platform was first implemented into a PCN in Leicester. HCP experiences were gathered through video call interviews at different stages of this implementation.

This service evaluation design consisted of three phases. Interviews were conducted with HCPs to gather their thoughts and experiences with the RocheDiabetes Care Platform at each phase.

### Study Design

The implementation of the RocheDiabetes Care Platform into the Leicester PCN involved HCP training, as well as onboarding a cohort of PwD onto the platform. HCPs attended a one-hour training session on the RocheDiabetes Care Platform and its functions. Following this, HCPs booked PwD into ‘onboarding appointments,’ where the mySugr app was downloaded onto the PwD’s phones. All PwD were instructed on the tool itself and the correct way to use it.

There were three stages to the service evaluation: pre-implementation, implementation, and post-implementation. Semi-structured interviews were conducted with 13 participants from the Leicester PCN. Interviews were conducted over a video call on Microsoft Teams; the interviews were video and audio recorded.

### Participants

Participants were from a variety of roles directly associated with diabetes care within the PCN (Table 1). Conducting interviews with a wide range of positions ensured that the impacts of the digital tool, across all areas of primary care, were captured.

**Table 1.** Roles of healthcare professionals participating in the service evaluation.

| Role within PCN | Number of participants |
| --- | --- |
| Lead pharmacist | 1 |
| Pharmacist | 2 |
| GP | 1 |
| GP with a specialism in diabetes | 1 |
| Practice nurse | 3 |
| Diabetes specialist nurse | 1 |
| IT lead | 1 |
| IT | 1 |
| Administrator | 1 |
| Business Manager | 1 |
| Practice manager | 1 |

### Data Collection

Data collection was performed independently by the Research Team at Nottingham Trent University. There were three opportunities for data collection throughout the service evaluation. The first interview was conducted in the pre-implementation stage, prior to the implementation of the Roche Diabetes Care tools. This interview examined the current management of the PCN’s PwD, including the tools employed for diabetes care. The second interview was conducted after the six-month implementation stage, as HCPs became accustomed to the tool. This interview explored the benefits and challenges that arose from the initial use of the tool. The third interview was conducted at the post-implementation stage, with the Lead Pharmacist, at the end of the service evaluation. The individual interview was necessary as the Lead Pharmacist had the most involvement with the tool and was the lead in managing the care for PwD. This interview explored the effect of the digital tool on personalised care for PwD, as well as individual outcomes. This interview also investigated the impacts of the digital tool on consultations and HCP workloads. All interviews were audio recorded and transcribed. The HCPs were asked about their demographic and background information at the beginning of each interview.

### Data Analysis

The data was analysed using Thematic Analysis. Wiltshire and Ronkainen’s^11^ realist approach to thematic analysis was followed during data analysis. Furthermore, the accuracy of transcripts was ensured through the transcription by two individual researchers.

## Results

Here, the results from the pre-implementation and post-implementation stage are presented. These results include the words of HCPs, as well as their answers to Likert scale and ranking questions. The themes that were found using thematic analysis are additionally presented in this section.

### The Pre-Implementation stage

Interviews were conducted prior to the implementation of the Roche tools, which identified the PCN’s pathway of care for PwD. This included challenges that the HCPs experienced. Themes were identified during this stage. Pre-appointment activities and appointment activities involved actions following the appointment. Themes included challenges that were raised by HCPs, presented below. Figure 6 illustrates the PCN’s pathway of care prior to the implementation of the Roche tools.

**Figure 6.**
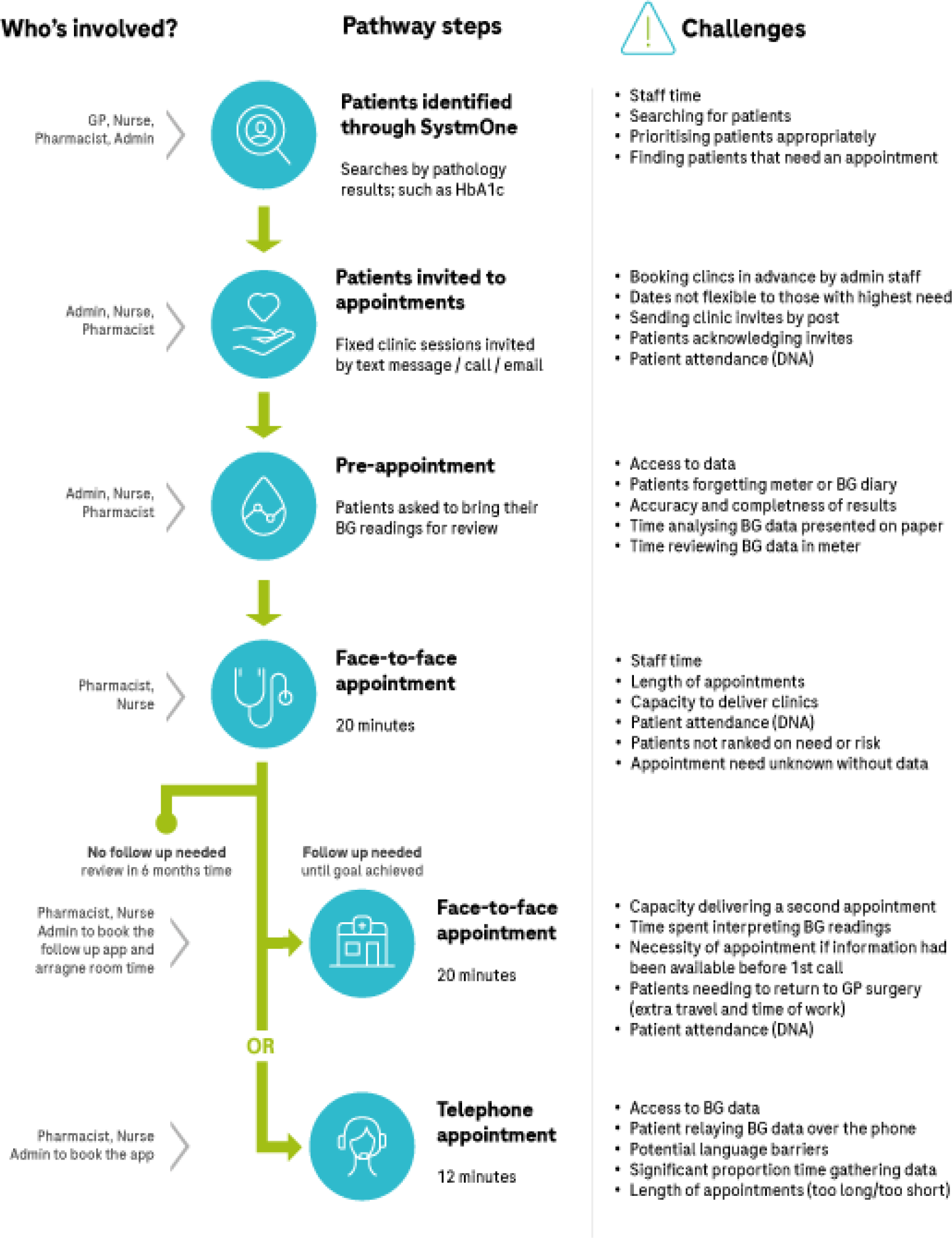
PCNs Pathway of Care for PwD, prior to implementation of the RocheDiabetes Care Platform.

#### The Pre-Appointment Stage

This stage involved searching for PwD who needed reviews, using searches and notes on a electronic medical record system. This electronic medical record was the main method of diabetes management for HCPs, prior to the implementation of the RocheDiabetes Care Platform. In this pre-appointment stage, HCPs would send invitations to PwD *via* various communication channels, such as text messages, letters, or phone calls, and reminding PwD to bring in their blood glucose readings. In the Leicester PCN, this responsibility was shared among administrative personnel, nurses, pharmacists, and GPs.

#### Challenges within Pre-Implementation

One of the main challenges, highlighted by HCPs, was finding PwD who required a face-to-face (F2F) consultation. HCPs found difficulty in this area due to the complexity of the electronic medical record searches, and the complexity of recorded patient data.

> *‘I’ve done my own search for people who’ve had an HbA1C in the last two years. Then going through…3,000 patients…to see did they have follow up bloods? What happened? Where was it? It’s a huge, huge task.’* - Practice Manager, Pre-Implementation Interview

> *“There’s not much in [the electronic medical record] that will support these patients going forward. Yes, there’s templates and you can go through those but in terms of something tailored a bit more to that condition, it’s quite exciting that there’s these apps coming out that will do that.”-* IT Lead, Pre-Implementation Interview

Furthermore, the pre-appointment stage was heavily managed by nurses; this involved identifying the PwD, and going through their records to determine who needed to be invited for an appointment or virtual appointment. This process was described as “*impractical*”(Diabetes Specialist Nurse, Pre-implementation Interview), especially when managing other nursing duties.

> *“Time needs to be set aside for all other nursing duties.”-* Diabetes Specialist Nurse, Pre-implementation Interview

> *‘In practice nursing, it’s quite a small team…I found this is the biggest challenge in this job.’ –* Practice nurse, Pre-Implementation The shortage of nurses was also highlighted as a challenge by HCPs. To address this challenge, HCPs called for a more *‘centralised*’ (Practice Manager, Pre-implementation) process that provided easy access to diabetes data, facilitating the identification of PwD who require F2F appointments.

#### The Appointment Stage

This stage consisted of GP, nurse and pharmacist consultations with PwD. In review appointments, PwD would have their height and weight measured and depending on the advice of the clinician, can have blood tests and foot examinations. A discussion with the Diabetes Specialist Nurse about the PwD and their diabetes plan would also take place. PwD would be asked to bring blood glucose diaries to these appointments.

#### Key Challenges in Appointment Stage

> *“Just the volume of diabetes that we’ve got and the complexity, I think and how limited we are with time and staffing.*” - Practice Nurse, Pre-Implementation Interview

> *‘[Patients] are usually asked to bring a urine sample or bring a record of their blood sugar recordings but some people forget or can’t find their book, it varies’* – Diabetes Specialist Nurse, Pre-Implementation Interview

> *“I would say the difficulties sometimes with the patient groups sometimes they’re not always responsive and don’t always want to come into the surgery for management or just lack of understanding on their part so it would be nice to have more facilities for the patients I think”* - Practice Nurse

> *’We had loads of information, but it meant nothing… The data points we had, but they really didn’t mean anything. We couldn’t interpret them because they were just all over the place.’ –* Lead Pharmacist on recorded diabetes data prior to implementation.

According to the Diabetes Specialist Nurse, the key challenges in the pre-appointment stage involved, *“getting patients to self-monitor, getting patients to submit their readings, not being reminded by staff to bring information when booking appt.”*

Key challenges in the appointments stage included, the lack of access to diabetes data, appointment attendance and limited time within the appointment for clinical decision making based on the data available. HCPs found it challenging to review blood glucose measurements due to a lack of diabetes and lifestyle data provided by the PwD. Although templates for diabetes were available on the electronic medical record, the data was not organised so that specific diabetes data could be found and patterns could be identified with ease. Instead, HCPs would take time scrolling through written data on the PwD’s record during the appointment. Sometimes, PwDs would forget to bring their blood glucose readings to appointments, leading to inefficient visits with limited progress. Moreover, HCPs explained that a lack of understanding and time for more in-depth communication may have contributed to PwDs forgetting to bring their blood glucose readings to appointments. This was a main challenge identified by the HCPs.

Working with PwDs to ensure blood glucose readings are available at appointments was highlighted as important for HCPs. However, there was an awareness that progress cannot be made with the current digital system’s lack of provision for managing diabetes.

HCPs noted that managing those PwDs who felt comfortable on a digital platform would allow for more time with those who preferred more traditional methods of communication, ensuring equality of care.

### The Post-Implementation Stage

Currently, the RocheDiabetes Care Platform is yielding tangible benefits in the Leicester PCN. This includes staff time and cost-savings, optimised appointments, enhanced engagement, and increased centralisation of resources.

There have been several changes since the implementation of the RocheDiabetes Care Platform. There were 29 PwD that actively used the mySugr app, with their diabetes data shared to the RocheDiabetes Care Platform. Previously, 21 of these PwD would have needed a F2F appointment. The pharmacist no longer requires a F2F appointment unless there is a major concern with a PwD based on the available data on the RocheDiabetes Care Platform. In addition, to complete the 21 F2F appointments, a clinic room would have been used for a full day at an internal cost to the practice of £240. This cost no longer exists and rooms are free to be used for other clinical activities.

### 6 months Post Implementation

#### Improved Glucose Control

“Most of the people that are pulling data [actively using the mySugr app] are improving on their standard deviation. So the standard deviation is flattening out, which means they’re in range a lot more. There’s a lot less fluctuation, a lot less hypo’s, a lot less hyper’s… It’s above the 60% of the people who are pulling data are improving their control.”

The Lead Pharmacist’s audit of the PwD who were onboarded onto the RocheDiabetes Care Platform showed that 17, of the 29 PwD actively using the digital tools, displayed improved glucose control.

#### Pre and Post-Implementation Comparison

The results presented below illustrate HCP experiences after a 6-month period of using the RocheDiabetes Care Platform. Therefore, these results highlight the initial thoughts and responses from HCPs about the use of the RocheDiabetes Care Platform and the potential short-term benefits it can deliver.

**Figure 7.**
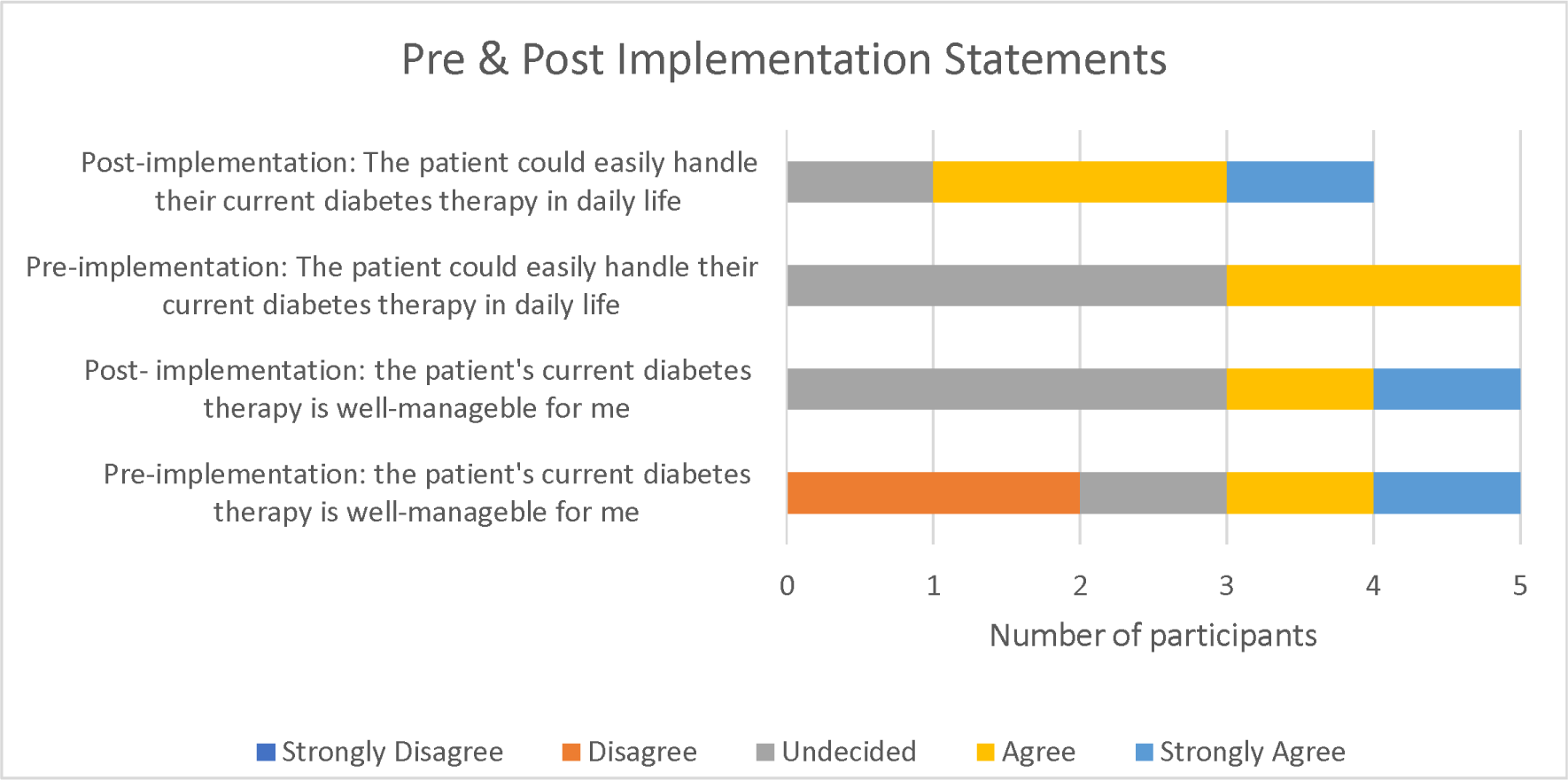
Likert Scale Results of Pre-implementation and Post-implementation Statements.

This graph includes data from the results of the implementation interview. Answers were provided by the lead pharmacist, 2 clinical pharmacists, the diabetes specialist nurse, and a practice nurse. These participants had interactions with PwD who were onboarded onto the RocheDiabetes Care Platform.

When questioned on the PwD and their ability to handle their diabetes therapy, an improvement was observed. This may be a result of the RocheDiabetes Care Platform functions for PwD, particularly with the instant sharing of recorded blood sugar readings. One of the participants preferred not to provide an answer to the post-implementation part of this question. Therefore, 2 participants agreed 1 participant strongly agreed with the post-implementation statement that ‘the patient could easily handle their current diabetes therapy in daily life’ and 1 participant was undecided.

Moreover, an improvement was identified in the HCP’s ability to manage their PwD, with two participants agreeing and strongly agreeing that the RocheDiabetes Care Platform had helped to improve their management of PwD.

This graph shows that 3 of the 5 participants who answered the Likert Scale questions, agreed or strongly agreed with the statement about the RocheDiabetes Care Platform’s benefits to the self-management of diabetes.

Figure 9 highlights that effective communication between HCP and the PwD, regarding blood glucose values has improved as a result of the RocheDiabetes Care Platform as all participants who were asked to answer the Likert Scale statements either agreed (2) or strongly agreed (3).

**Figure 9:**
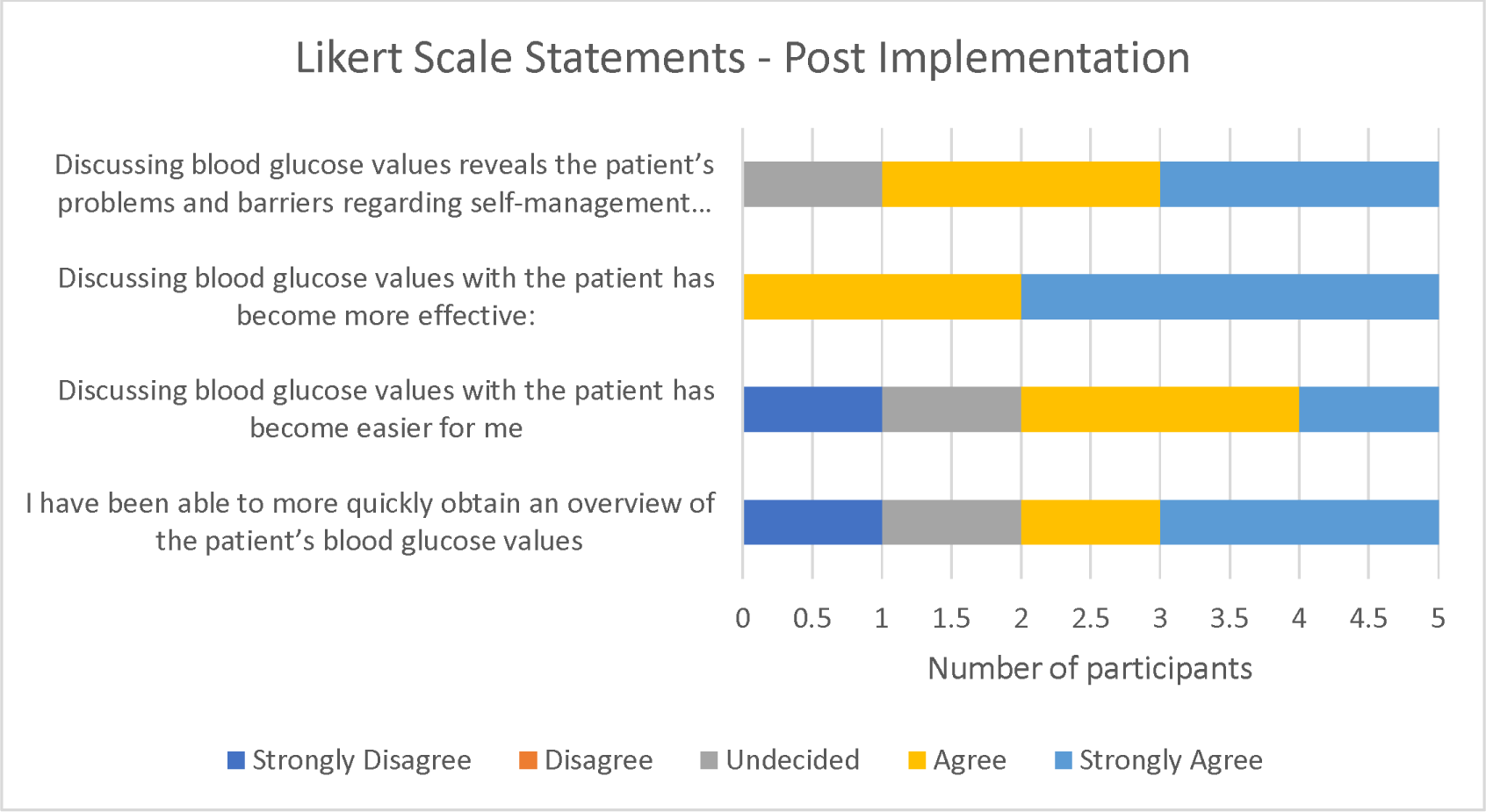
Post-Implementation Likert Scale Statements.

One participant strongly disagreed with the statement about the RocheDiabetes Care Platform ability to ease discussions about blood glucose readings with PwD. However, 3 of the participants either agreed or strongly agreed with this, with one participant undecided.

One participant strongly disagreed with the statement regarding overviews of patient blood glucose values. However, 1 participant agreed and 2 participants strongly agreed with this statement. One participant was undecided.

**Figure 10:**
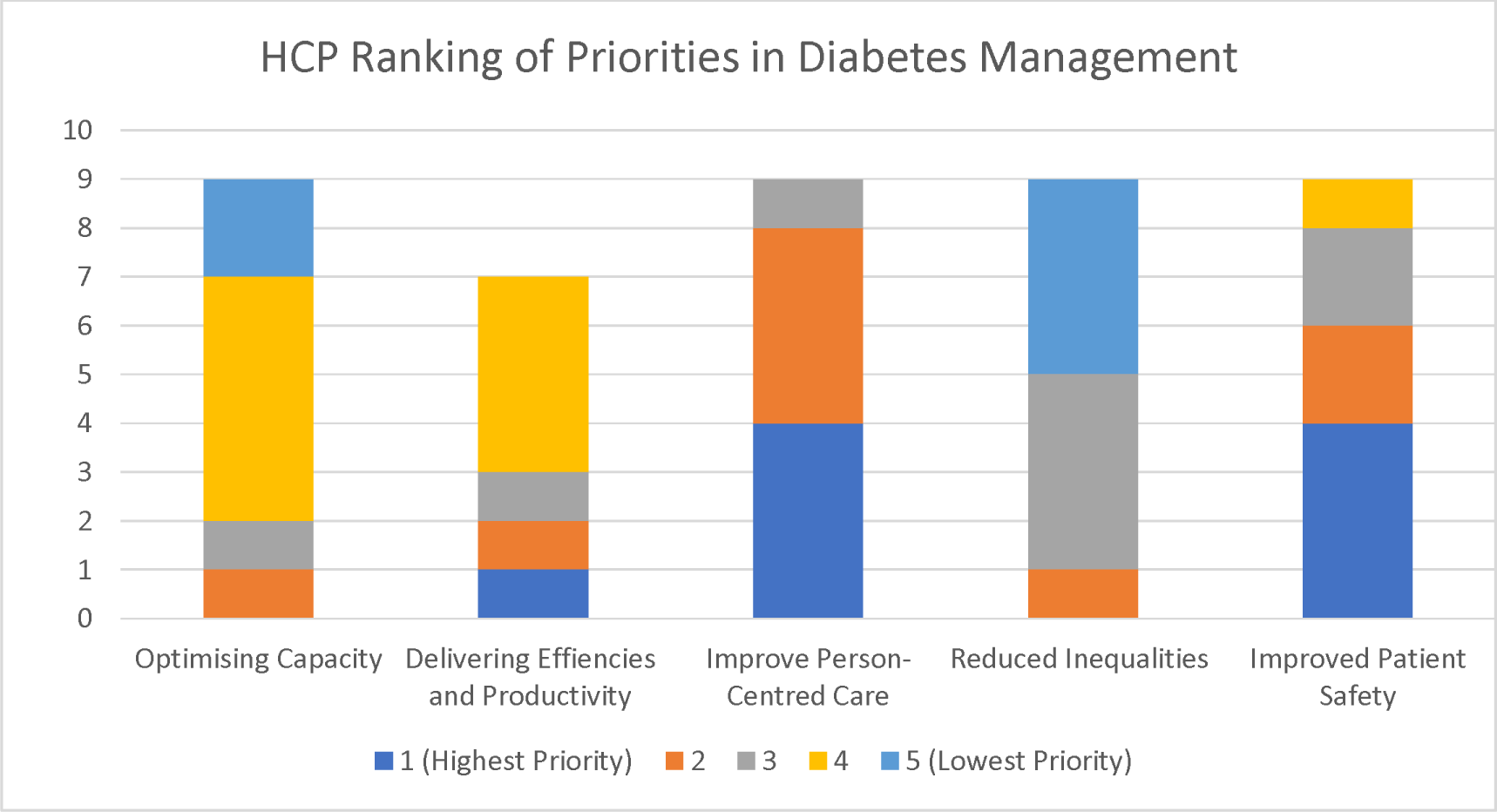
HCP Ranking Priorities Post-Implementation Interview Question.

HCPs were asked to rank priorities within diabetes management. Improving person-centred care and improving patient safety were shown to be the highest priority of participants. 4 of the 9 participants viewed improving person-centred care as their highest priority, and another 4 participants viewed this as their second highest priority.

Improving patient safety and care was more important to many of the participants than optimising capacity and delivering efficiencies and productivity, suggesting that HCPs are more concerned about the outcomes and well-being of their PwD. Reducing inequalities was perceived as the lowest priority by 4 participants.

#### Post-Implementation Interview Themes

Four themes were identified during data analysis, including impact on appointments, patient education and communication, centralisation of resources and future implications for the RocheDiabetes Care Platform. The themes are described below.

#### RDCP’s Optimisation of Appointments

> *“[Patients] don’t need to give me the readings anymore. They don’t need to find the book… when I ring, I ring with the agenda to hand. I’m not ringing to say, let me find out what the problem is, then do the review. I know the problem.” –* Lead Pharmacist, 12 months Post-Implementation Interview

> *“It [the* RocheDiabetes Care Platform *] allows us to…look at the results maybe prior to the appointment saves time that way and then at least as a clinician we have a bit of a better plan about what we’re going to do with that patient.*” – Pharmacist, 6 months Post Implementation Interview

The RocheDiabetes Care Platform has the potential to reduce appointment duration and effectively resolve the issues of data retrieval, previously faced by HCPs. HCPs no longer need to spend time interpreting or investigating recorded blood glucose values in appointments. This enables HCPs to have valuable, data-driven conversations with PwD, allowing them to begin consultations with a pre-established agenda instead of having to formulate one during the appointment. This may reduce both the preparation time and overall duration of appointments. Previously, PwD often forgot to bring their blood glucose readings, leading to unproductive appointments, however, this is no longer a challenge.

#### Reduction in F2F Appointments

> *“Now there’s no patient F2F reviews unless necessary. If I have seen them once before, I probably don’t need to see them again for a face to face. All the data I need is available digitally now.”*

> *“The data now is irrefutable.”*

> *“Back in the day, you’d take annual leave or don’t go in or work from home, whatever it might be to come and see me just for half an hour appointment. Now you aren’t taking any time off…you can take the call at work!”*

- Lead Pharmacist, 12 months Post-Implementation Interview

> *“[Patients] often don’t bring their book in for me to look at their results, but on the screen I can see it”*. - Practice Nurse 6 months Post-Implementation Interview

> *“I’ve been able to be more direct with the patient because obviously…I know when they come and see me, they’re not going to tell me the truth, you know, because they want the diabetic nurse to be nice to them. And this is what she wants to hear. Well, it’s not, I want them to be honest. So I can help them.”* Practice Nurse describing the difference in monitoring blood sugar readings using the RocheDiabetes Care Platform, 6 Months Post-Implementation

Since onboarding onto the RocheDiabetes Care Platform, there have not been any F2F appointments with the PwD who are actively using the mySugr app unless there were major concerns. The implementation of the RocheDiabetes Care Platform can allow for more time with PwD who need additional support, as well as facilitating a remote way of working that accommodates both HCP and the PwD. The previous reliance on F2F appointments was caused by a lack of consistency in the recording of diabetes data, such as blood glucose readings. HCPs felt the need to examine their PwD to ensure these values were reliable. Often appointments consisted of going through PwD’s diaries and deciphering values, which had been recorded with the wrong date or time. However, HCPs can now rely on the RocheDiabetes Care Platform,’s comprehensive display of diabetes data.

Accuracy in blood glucose readings is important for HCPs, particularly when making decisions about the PwD’s plan. The combined use of the Roche tools for diabetes may increase accuracy for clinicians and simultaneously empower PwD to confidently self-manage their diabetes.

The reduction in F2F appointments is particularly beneficial to PwD who often face difficulty in scheduling appointments due to their work commitments. The increase in remote appointments is efficient and accommodating as PwD can attend appointments from any location.

#### Reduction in Frequency of Appointments

> *“The appointments of the people that I don’t need to see have definitely dropped.”*

> *“Those who are really well-controlled will just get a text message from us once or twice a year.”* Lead Pharmacist, 12 months Post-Implementation Interview

Here, the lead pharmacist observed a reduction in the frequency of both F2F and remote appointments after the implementation of the RocheDiabetes Care Platform, saving both HCP and PwD time. Previously, it was difficult for HCPs to discern whether an appointment was always necessary, as highlighted in the quote below from the Lead Pharmacist, describing the pre-implementation challenges.

> *“I don’t know you’re well controlled until you come into my door because I’ve not looked at your data. Then, you are in and out within 5 minutes. But it’s taken 20 minutes of my time.”-* Lead Pharmacist, Pre-implementation.

Sometimes, PwD who were achieving their targets and did not require a face to face appointment were taking time to attend the clinic instead of being reviewed over the phone. The RocheDiabetes Care Platform’s accessible presentation of diabetes data can allow HCPs to quickly identify the PwD that do require appointments, potentially reducing the overall frequency of F2F visits. Thus, PwD may be able to save time and money by attending appointments remotely.

##### Reduction in Did Not Attends (DNAs) in Appointments

Since the implementation of the RocheDiabetes Care Platform, there have been 0 DNAs from PwD who had been actively using the mySugr app. The RocheDiabetes Care Platform’s facilitation of remote visits and accommodation for PwD’s lifestyle may reduce the number of DNA appointments. Appointments are more flexible; if a PwD misses their telephone consultation, the HCP can send a text message offering other suitable times. This indicates that RocheDiabetes Care Platform can significantly expand the options available to HCPs to work with PwD’s lifestyles.

> *“I’ll send a patient a message [stating that] I just called from the surgery to discuss your sugar reading I’ve looked at on online. Let me know when is suitable for you to discuss these so then we can agree a suitable time between us.”*

> *“So, if a patient doesn’t pick up, it’s not a DNA. It’s just an opportunity for another patient to have their spot instead. It’s no longer wasted. I’m not sitting for about 10 minutes for the next patient. Now I just call the next patient up…I’m not wasting my time.”*

- Lead Pharmacist, 12 months Post-Implementation Interview

Previously, HCPs wasted significant time on DNAs as they had to wait for their next appointment. Further resources would have been utilised, such as organising admin to re-book the appointment, and booking another clinic room to see the PwD. Now, the repercussions of a DNA are lower; the HCP can make use of their time and communicate new consultation times with the PwD easily.

#### Time and Cost Savings in Appointments

> *“A GP costs £100 P/Hr. Add in 20% on costs = £120.”- Business Manager*

> *“A Pharmacist costs £40P/hr. Add in 20% on costs = £48/ph”* - Business Manager, Post-Implementation Interview

When asked how many PwD were reviewed in an hour prior [to RDCP], the Lead Pharmacist answered *‘3.’* When asked how many PwD were reviewed in an hour after RDCP, Lead Pharmacist answered, *‘up to 10.’*

The above results indicate the time and cost savings in appointments as a result of implementing tools such as the RocheDiabetes Care Platform. The Lead Pharmacist indicated that the platform may allow for more PwD to be seen within an hour due to the time saved in appointments with PwD.

The additional 20% of costs in the Business Manager’s statement consisted of the clinic room cost. This cost no longer exists, as the Lead Pharmacist stated that there have been no F2F appointments required since the implementation of the RocheDiabetes Care Platform, into the PCN.

#### Enhanced Engagement with PwD

Following the implementation of the tools into the PCN, HCPs observe an improvement in engagement and interactions with PwD. The RocheDiabetes Care Platform works well for individual PwD. For example, PwD who take an avid interest in their condition now have the opportunity for more oversight and control of their management. The mySugr app use of graphs and visual data aim to deepen a PwD’s level of understanding about diabetes. HCPs also describe the way in which the mySugr app may work to encourage engagement for PwD who struggle to engage with diabetes management. The RocheDiabetes Care tools’ advances in blood glucose monitoring, coupled with the uncomplicated display of diabetes data, underscores the RocheDiabetes Care Platform as a mechanism that may motivate PwD to participate in the management of their condition.

> *“We have patients who are quite invested in their own therapy… we introduced [the] new technology to them which might even further improve their…management, because they’ll be further invested”*

> *“For those patients we couldn’t get to engage with us…what we did was say this is a new tool we have at our disposal to try and get them engaging with us…saying look, we’ve got something that’s new, it’s a new bit of technology, let’s try it out!”*

- Nurse 6 months Post-Implementation Interview

> *“At the end of the day, [patients] need to make those changes and obviously, as a clinician it is important, we give them the information so they can make that informed decision… I think it just comes down to discipline and… I suppose, giving them the tools like the Roche app … where they have more control and more greater understanding of the blood sugar.”-* Pharmacist, 6 months Post Implementation

Moreover, HCPs recognise the RocheDiabetes Care Platform’s ability to potentially educate PwD further; the tool alerts HCPs when a PwD’s blood glucose reading is out of target range. PwD can also see the trends in their blood sugar readings, allowing them to see the benefits of improving diet, lifestyle, and medication.

> *“It’s got the colour coded graph on the side… patients really like that.”- Diabetes Specialist Nurse-6 months Post Implementation Interview*.

**Participant 2 Case Study: Post-Implementation of RDCP**

**Case study of PwD described by Lead pharmacist in PCN**

Participant 2 has Type 2 Diabetes and previously found difficulty when performing certain lifestyle activities. Participant 2 would had spent much time in appointments going through different blood glucose reading with the Pharmacist.

Participant 2 is now onboarded to RDCP; the pharmacist can clearly see patterns in Participant 2’s blood glucose readings. RDCP highlighted that Participant 2 experienced low blood sugar every [specified day of week].

The Pharmacist and Participant 2 discussed activities the patient performs on [specified day of week]. Pharmacist learnt that the participant went on a long walk, which is also when they experienced particularly low blood sugar.

The Pharmacist was able to provide informed guidance to Participant 2, advising them to eat before the [specified day of week] walk. RDCP allowed pharmacist to identify the problem with ease, communicate with the participant and adjust the participant’s lifestyle in one appointment.

Participant 2 is now able to go on their [specified day of week] walk without experiencing low blood sugar levels.

Pharmacist stated that without the RocheDiabetes Care Platform, ‘I’d never have been able to find that [specified day of week] irregularity and give that advice, specific to that day.”

“I don’t need to spend 20 minutes asking where [Participant 2] is [the rest of the week]. That. Doesn’t bother me. It’s what he does on [specified day of week] because that’s where the problems are.”

The mySugr app can provide accessible diabetes data, accessible to the PwD at all times. In the interviews, HCPs stress the importance of family involvement in healthcare for some PwD, particularly when managing PwD from diverse cultures with diverse traditional values and needs. HCPs describe using the RocheDiabetes Care Platform during family interventions. For example, one HCP onboarded a PwD in their 70s. Despite concerns that the digital tool would be too complex for the PwD, the RocheDiabetes Care Platform accommodates the PwD’s comforts and needs, such as family involvement.

With the PwD’s consent, the tool was set up on a family member’s phone, enabling them to assist with uploading blood sugar readings, and communicating vital information to the PwD. A diabetes management solution which can support a wider conversation between PwDs their families and their HCPs such as RDCP and mySugr could therefore provide an additional benefit in self-management with family support.

> *“Overall, we’ve improved health inequalities… because not only is the patient getting what she was before anyway…but now the daughter can keep an oversight and we have also got access [to patient’s data]. So that has improved it!”*

> *“On RDCP, you’ve got full access as soon as the patient tests their blood sugar…I can see that the patient had tested. I can see the patients who haven’t tested, and I can send them a message.”*

- Lead Pharmacist, 12 months Post-Implementation Interview

**Figure 11.**
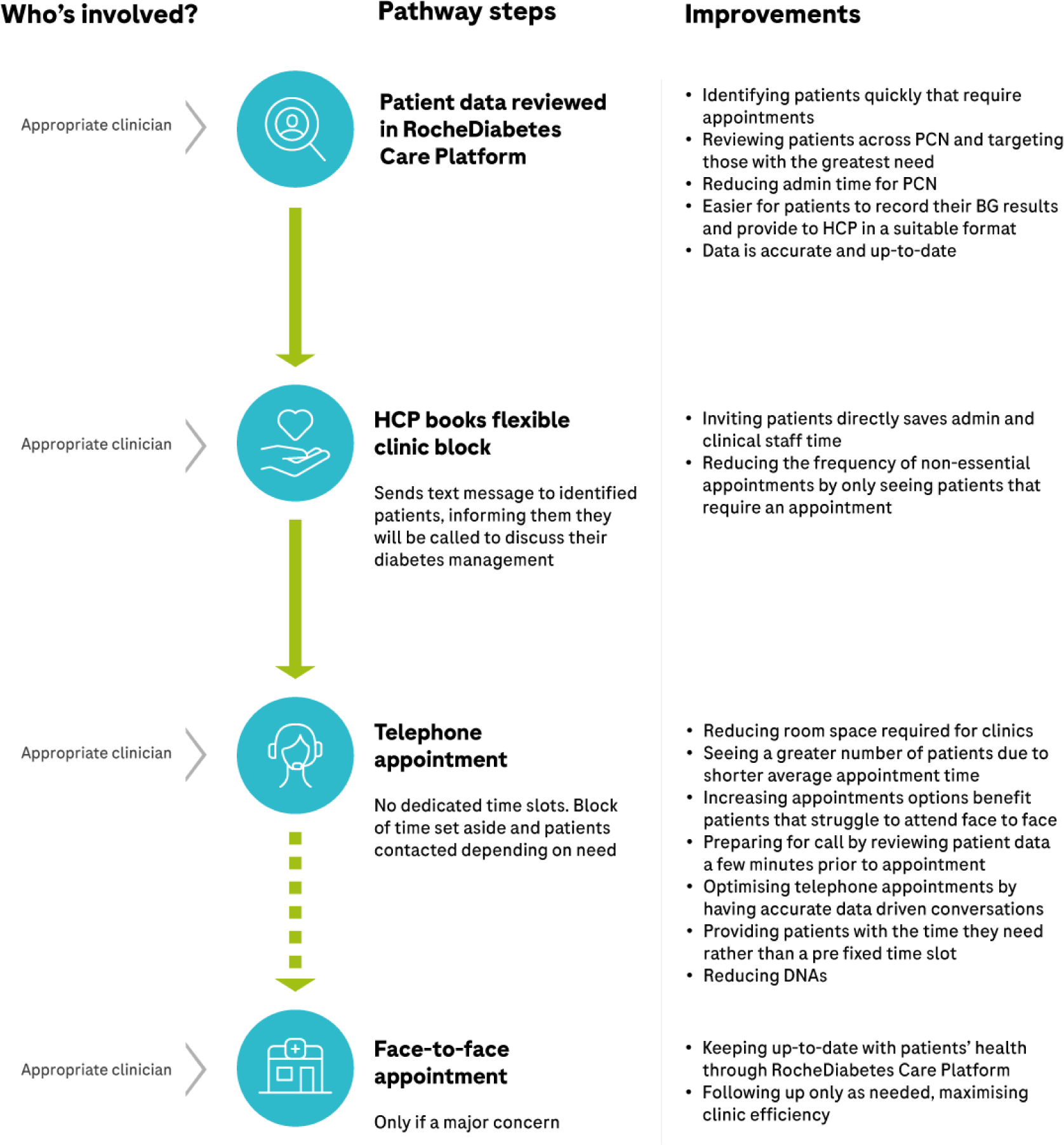
Modified pathway following implementation of the RocheDiabetes Care Platform.

Additionally, HCPs acknowledge that the RocheDiabetes Care Platform could have helped to reduce the damaging effects of the pandemic on diabetes management. The functions of the platform and the app could have alerted HCPs to PwD who were struggling to manage their diabetes, providing an efficient way to identify PwD who required telephone consultations. The RocheDiabetes Care Platform’s function of highlighting HbA1c levels that are out of range, may have increased PwD’s understanding. One HCP recounts the challenge of patients being unable to discern dangerous HbA1c levels. The RocheDiabetes Care Platform display of clear and visual diabetes data may have allowed HCPs to identify and communicate with PwD who needed support during the pandemic.

#### Centralisation of Resources

The Leicester PCN were working to merge GP sites during this service evaluation. To ensure a seamless transition during the merger, centralising resources was crucial for the PCN. Many of the HCPs believe that the RocheDiabetes Care Platform can improve the centralisation of resources by providing a simple and consistent digital platform to manage PwD across all GP sites. All HCPs see the same diabetes data, and no longer need to use a variety of platforms to interpret this data.

> *“It’s tools like this that make [people] more efficient… with a template you just end up typing in different texts and fields and there would be no organisation to that in the future, it wouldn’t be very easy for someone to understand what you’ve done. Anything that is more consistent, I’m up for.”*

> *“It’s good to have prompts, especially for folks who are new to that particular clinical area or are maybe not as experienced as someone who has got many years of experience.”*

– IT Lead 6 months Post Implementation

RDCP’s ability to centralise resources also provides the opportunity for the upskilling of staff. Some HCPs explain the training session provided by Roche Diabetes Care and commend the way these sessions were understandable for all roles within the primary care network. The digital platform and its training are described as ‘straightforward.’ The business manager in the Leicester PCN emphasises the digital tool’s ability to further utilise their staff as the RocheDiabetes Care Platform’s uncomplicated display of diabetes data also benefits HCPs who are new to the clinical area of diabetes care.

Moreover, the RocheDiabetes Care Platform also benefits PCNs with limited staff and resources. For example, a HCP with specialism in diabetes now has remote access to data from the PCN’s entire population of PwD. The platform has the ability to make tasks such as altering PwD’s plans, or changing titration of insulin, more efficient.

> *“Across our network we’ve only got a few insulin specialists, and we’ve got quite a lot of patients on insulin. So, if the nurse at one practice has access to the patient [who is] somewhere else, we have this centralised resource. We can deal with these patients because we’ve got the information to hand. The patient can be at home, and we’ve still got remote access to all of this.”*

- Lead Pharmacist, 12 months Post-Implementation Interview

#### Pathway of Care for Diabetes Review Appointments

1. HCP uses the RocheDiabetes Care Platform to identify PwD who need a consultation.
2. HCP books time for remote clinics
3. HCP sends text messages to identified PwD, informing them that they will be called to discuss their diabetes management.
4. HCP can prepare for a call by reviewing the PwD on the RocheDiabetes Care Platform a few minutes prior to the appointment.
5. HCP consults with the PwD – (approx. 6 minutes per patient)
6. HCP can keep up to date with the PwD’s health through the RocheDiabetes Care Platform and organise another review if necessary.

* Pathway of care described by the Lead Pharmacist in the PCN.

#### Challenges in the Old Pathway of Care that the RocheDiabetes Care Platform has Addressed

Following the implementation of the RocheDiabetes Care Platform, the pathway of care for diabetes is streamlined, eliminating previous challenges for HCPs, such as the difficulty in identifying patients who require an appointment, the lengthy patient invitation process, and the reliance on patient-recorded data.

One HCP labeled RDCP as a “population health management facilitator,” explaining the benefit in RDCP’s display of data and trends, which allows the HCP to easily identify PwD that require an appointment.

##### “The people who need my time are getting my time”

To an extent, the pathway remains the same for newly diagnosed PwD, who may still need to come in for height/weight, foot, and blood tests. The change of the patient pathway is indicated in review appointments. There is no longer the need for an appointment with a nurse to check if the PwD is recording their blood glucose, as this data is available for HCPs digitally. The duration of appointments is also shorter.

Additionally, the RocheDiabetes Care Platform works to supports different ways of working for HCPs. One HCP can use the digital platform to accommodate their own method of managing PwD. For example, they no longer need to book in clinic days. The platform’s access to diabetes data enables the HCP to complete necessary appointments whenever at a time which suits them. Whilst the tool is still utilised by other HCPs who prefer clinics, the use of the RocheDiabetes Care Platform allows for a more flexible style of working for this HCP.

The Onboarding Process

> *‘It’s just mainly the onboarding of patients. I think that just takes up a lot of clinical time, which is why when we started it, I was very clear in saying that we as clinicians, we don’t really have the time to onboard patients. So we needed the kind of Roche’ expertise and input.*’ – Pharmacist, 6 months Post Implementation

> *“I would say the time that it takes and how savvy your patient is with their tech as well because it is quite a lengthy process and if they’re not great with a mobile phone, it’s quite hard work.”* Practice Nurse on challenges with onboarding process, 6 months, Post-Implementation.

HCPs identify challenges with the initial onboarding process, where PwD were taught how to download the app and the app was synced to the RocheDiabetes Care Platform. The HCPs explain that the process could be quite lengthy, due to a number of reasons, including the patient’s phone not being charged, internet connections and the extra time for explaining the new app and digital tool to the PwD. One HCP said that they benefited from the company’s involvement in the onboarding process, especially as the HCPs adapted to the RocheDiabetes Care Platform themselves.

### 5. Future Vision of PCN with RDCP

> *“I don’t see why anybody would be on…non-digital forms of measuring. Everybody should be given the opportunity to do this.”*

> *“It’s allowed me to do all these sorts of things. I’ve never had that chance on the manual way…I do it safely at the same time protecting my registration and my clinical ability to survive and this allows me to do that.”*

In this PCN, the Lead Pharmacist maintains an integral role in decisions on the investment of digital tools for diabetes. The lead pharmacist has a future vision of the RocheDiabetes Care Platform and their PCN, describing their aim to onboard the PCN’s entire cohort of PwD onto the RocheDiabetes Care Platform. The duration of the current onboarding process is a barrier at this point in time and has been highlighted by many other HCPs. However, the Lead Pharmacist believes that the RocheDiabetes Care Platform is a more secure way of managing PwD

The Lead Pharmacist expands on the tools use of visual graphs, which transforms data into something ‘digestible’. With the clear trends of standard deviation and variation, the Lead Pharmacist finds it easier to identify problems and develop solutions for PwD who fall out of range. The HCP explains that the digital tool transcends diabetes management, suggesting that the RocheDiabetes Care Platform not only helps to manage blood glucose control, but also expands diabetes management to encompass other health factors.

## Discussion

The results of this service evaluation highlight the potential for the RocheDiabetes Care Platform to support iPDM by potentially improving the communication between HCPs and PwD. There are several opportunities for improving the management of PwD in primary care through the implementation of the RocheDiabetes Care Platform, most poignantly within the improvements in staff time and reducing costs, as well as PwD’s engagement.

### Staff Time Saving and Cost Savings

The RocheDiabetes Care Platform, has presented the opportunity for time and cost savings as a result of the reduction in F2F appointments and the increased number of PwD reviewed per hour. Since there have not been any F2F appointments since the implementation of the RocheDiabetes Care Platform, the hypothetical time and cost savings have been calculated.

During this evaluation 21 appointments with PwD that would have normally been held in the surgery were avoided. The pharmacist’s time, at a cost of £48/hour, equates to a saving of £336 over a 7 hour working day. With the additional cost of a clinic room at £240 prevented, this comes to a total cost saving of £576. Typically, a PwD in this PCN will have 3 appointments a year with potential cost saving for this cohort of the RocheDiabetes Care Platform, onboarded PwD of £1,728. The Leicester PCN currently has 3,127 PwD. Onboarding this cohort of PwD onto the RocheDiabetes Care Platform, could result in the approximate cost saving of £257,299 a year.

### Optimised Appointments

The implementation of the RocheDiabetes Care Platform helped to alleviate some of the challenges identified in the pre-implementation phase by the HCPs, allowing for more optimised appointments. For example, in the pre-implementation interview HCPs described the challenge with recorded blood sugar readings as readings may be inaccurate or simply missing. However, the post-implementation results show that many HCPs have experienced an improvement after using the mySugr app and the RocheDiabetes Care Platform. The digital app worked to provide PwD with a way to record their blood sugar results using a mobile phone, with direct connection with the RocheDiabetes Care Platform. This has allowed for improved communication with PwD and optimised appointments as well as the ability to have improved personalised care. Person-centred care was shown to be a high priority for the clinical HCPs in the post-implementation interview. The case study and HCP experiences presented in this paper further underline the opportunities for personalised care as a result of the RocheDiabetes Care Platform as well as the ability for both PwD and HCPs to discuss individual challenges and goals. However, to further evaluate the impact of using the digital tools on patient reported outcomes, it may be useful to conduct interviews with the PwD who were onboarded onto RDCP in this service evaluation. Moreover, HCPs ability to discuss individual targets with PwD, as a result of the RocheDiabetes Care Platform, is important within diabetes management as individualised care can reduce the risk of complications from diabetes.^12^ The results from the service evaluation highlight that using the RocheDiabetes Care Platform can provide HCPs with increased confidence in the data reported by PwD, potentially reducing difficulties surrounding interactions in appointments, and encouraging constructive discussions with HCPs about PwD’s individual goals.

### Enhanced Patient Education and Engagement

Interviews with the lead pharmacist revealed that a combined use of the RocheDiabetes Care Platform and the mySugr app may increase PwD’s engagement with their self-management of diabetes. The additional digital support may have increased the meaningfulness of blood glucose monitoring.Research suggests that education and engagement with self-blood glucose monitoring practices improve self-management of diabetes. Furthermore, improvements in these self-care behaviours correlate with improvements in PwD’s quality of life, as well as better glycaemic control.^13^ Whilst the RocheDiabetes Care Platform’s potential benefits for PwD is described by HCPs in this service evaluation, research has called for more patient-centred outcomes in research on diabetes.^14^ Therefore, further research on PwD and their experiences with the mySugr app and the RocheDiabetes Care Platform is required.

This service evaluation highlighted the importance of patient engagement in the adoption of digital tools in healthcare. The PCN initially onboarded 63 PwD onto the RocheDiabetes Care Platform, at the end of the evaluation there were 29 active users. This translates to a 46% continuation rate in this study, meaning that 46% of PwD onboarded onto the RocheDiabetes Care Platform, have continued to record data for the 12 month duration of the PROVIDE study and after. Previous studies report similar continuation rates for digital health interventions. In a systematic review on drop out rates of digital health interventions in skin cancer, there was a median of 43% in study completion rates for observational studies.^15^

There were PwD who did not continue to record data for the 12 month service evaluation, understanding the reasons for this is an important element to further successful implementation. This may have been due to a lack of communication to PwD, regarding the benefits of the digital tool, as well as a lack of time for HCPs when implementing the onboarding process. According to Carlford *et al.*, contextual factors, such as lack of time, due to larger workloads, can affect a PCN’s ability to adopt new innovations^16^. Therefore, the challenge with the onboarding appointment may have also stemmed from the HCP’s coinciding pressures and responsibilities. Further research needs to be done to understand how the initial onboarding appointment can be improved for PwD as patient engagement in the adoption of new tools in healthcare is vital and can affect the success of a service redesign.^17^

### Streamlining Pathways and Centralising Resources

Since its implementation, using the RocheDiabetes Care Platform has helped to streamline the PCN’s pathway of care for PwD, saving HCP time in various ways. The implementation of the digital platform may have helped to mitigate the common complexities of streamlining pathways of care in health, that Rizan *et al* explains can result in ‘early abandonment,’ of newly proposed pathways of care,^18^ the RocheDiabetes Care Platform allows for the use of Rizan *et al*’s key steps for streamlining pathways of care. This includes identifying problems with the patient pathway, identifying the potential to streamline, forecasting the benefits of the streamlined pathway, planning the practicalities, and implementing and monitoring the streamlined pathway. This service evaluation has illustrated the potential benefits and opportunities for improvement with the RocheDiabetes Care Platform, as well as a method to implement the platform into primary care. This includes assessing the experiences and challenges at each stage of implementation.

The implementation of the RocheDiabetes Care Platform into the PCN has emphasised the value that lies within digital tools in healthcare. The service evaluation explored how access to digital patient data optimised appointments and allowed HCPs to complete diabetes tasks from various locations. This means that HCPs can access patient data from any site or surgery. This can be beneficial for HCPs especially during periods of staff shortage and in events when staff members need to limit their travel between the sites. This allows for several advantages such as support with the monitoring of people with diabetes and providing necessary, on-time treatment when required. More in depth analysis of onboarded PwD’s HbA1C levels from pre-implementation to post-implementation may provide more insight into the potentially improved patient outcomes as a result of the RocheDiabetes Care Platform.

Moreover, the experiences of the HCPs indicate that RDCP may fulfil the aim of digital technologies, outlined by Kerr *et al*, including providing assistance in the monitoring and treatment of diabetes.^19^ The digital platform encourages the centralisation of resources by providing a platform which prioritises the continuity of recorded diabetes data, therefore HCPs can have access to a clear overview of PwD’s diabetes journey. The platform’s ability to highlight trends and abnormalities in blood sugar readings can provide support to HCPs who are new to diabetes care, facilitating the upskilling of staff, which is often a barrier to the adoption of new tools due to training requirements. The HCPs involved in this evaluation reported that training was simple and straightforward and the tool offers benefits to HCPs who are new to the area of diabetes.

### Conclusion

This service evaluation was performed in a Leicester PCN where the RocheDiabetes Care Platform and the mySugr app were implemented across 9 GP sites. The results showed that using the RocheDiabetes Care Platform can enable the transformation of a number of areas of the PCN’s management of PwD. The opportunities presented by remotely managing PwD were identified from the perspectives of several stakeholders. Since the implementation of the digital platform and app, the PCN has the ability to re-evaluate and re-design workflow around a digital ecosystem of solutions. This has many benefits, particularly in the freeing up of costly room resources, increase in PwD seen per hour, a reduction in the frequency of non-essential visits and development of a leaner more efficient service. The challenges in recruitment of suitably skilled staff can be met by leveraging existing staff time more effectively and potential for ‘left shift’ of tasks to less experienced staff members.

One of the key challenges in digital services is adoption by clinicians, systems and PwD. This paper illustrates some key aspects of how this can be achieved at a very practical level. The improvement in communication and reduction in the number of F2F interactions is clear, as well as an improvement of blood sugar control in 60% of this sample. These outcomes are likely to be significantly valuable to PwD, and an aspect of this deployment that will be explored in the future.

## Data Availability

All data produced in the present study are available upon reasonable request to the authors

## Acknowledgments

Roche Diabetes Care co-authored this paper. mySugr and Accu-Chek are trademarks of Roche. All other product names and trademarks are the property of their respective owners

